# Evaluation of computer vision syndrome in the sample of students from the College of Medicine/University of Kerbala result of online learning with the isolation of COVID-19

**DOI:** 10.1101/2023.09.25.23295828

**Authors:** Mahdy H AbuRagheif

## Abstract

**Background:** A complex of vision problems related to stress the vision is experienced during the use of the computer or any digital device. Many of the visual problems that users report is just transient and go away after they stop using computers or other digital devices. Even after stopping their computer job, some people may still have diminished visual abilities, such as blurry distance vision.

**Subject and Method:** This is a cross-sectional study conducted at the University of Kerbala College of Medicine, which was completed between November 2021 and January 2022, to students of the College of Medicine given student questionnaires about the effects of long-time use of computers, laptops, and mobile phones.

**Result:** The cross-section studies with questionnaires to 460 students of the College of the Medicine/ University of Karbala, included these studies, we drew lines for the criteria to identify a computer vision syndrome from these associated ocular and extraocular symptoms and recorded a high prevalence of the Computer Vision syndrome reached 91.6%.

## Introduction

The development of the 21st century resulted in the educational sector of universities having a drastic change in the use of computers in learning. In addition, computers are used for watching movies, playing computer games, and online chatting. In early 2019 Iraq entered a political crisis in which enrollment in colleges and schools was prohibited, and after that came the COVID-19 crisis, which lasted for more than two years and continued. The method of teaching has become online in schools and universities through online teaching programs and social media As a result of using mobile phones, iPads, and computers for long hours, recorded harm to students of the College of Medicine/University of Karbala other than infection with COVID19, and in the last two years, the dependence on mobile and computer devices became largely due to the spread of COVID-19, which obligates many universities and schools to use audio-visual equipment for online learning and research work lead to complication of ocular and non-ocular sign and symptom same as computer vision syndrome in this research explained this syndrome in student. (1, 5, 9). Prolonged use of computers and other digital electronic devices often leads to a group of symptoms and an increase in the number of patients complaining about ocular and non-ocular symptoms related to computer use. Eye-strain, tired eyes, irritation, burning sensations, redness of eyes, dry eyes, blurred, and double vision reported by the visual display unit users was termed “Computer Vision Syndrome” (2, 5, 8, 9)

### Computer Vision Syndrome

A complex of vision problems related to stress the vision is experienced during the use of the computer, or any digital device “according to the American Optometric Association (3, 4). Many of the visual problems that users report are just transient and go away after they stop using computers or other digital devices. Even after stopping their computer job, some people may still have diminished visual abilities, such as blurry distance vision. If the root of the issue is not addressed, the symptoms will persist and may get worse with further usage of digital screens. (5,6) The degree to which people are affected by visual symptoms frequently relies on their level of visual ability and the length of time they spend staring at a digital screen. Inadequate eye focusing or eye coordination abilities, uncorrected vision issues like farsightedness and astigmatism, and aging eye changes like presbyopia can all lead to the emergence of visual discomfort when using a computer or digital screen device. (5, 6, 7, 8). Health issues with mobile phones include headaches, sleep disturbances, loss of concentration, short-term memory impairment, dizziness, burning skin, high blood pressure, glare from a digital screen, poor lighting, insufficient viewing distances, bad posture while seated, uncorrected vision issues, or a combination of these factors may be to blame for these symptoms [2]. These problems reported by the visual display unit users were termed “computer vision syndrome”[8,11,13].

### Risk factors

Multiple factors cause Computer vision syndrome; Numerous causes and treatment options have been identified in relation to symptoms. The course of treatment must be customized for each patient. To identify areas where our knowledge of the issue is lacking, nevertheless, a substantial body of research is still needed. In order to reduce productivity loss and morbidity from the condition, a specially created ocular inspection for computer users and concomitant counseling about the current best practices in computer use would go a long way., A few crucial elements in preventing or minimizing the symptoms of computer vision syndrome have to do with the computer and how it is used, including proper body alignment for computer use. This covers the ambient lighting, the comfort of the chair, the location of the reference materials, the placement of the monitor, and the use of breaks.

#### Sleep and circadian rhythm disturbance

The evening hours immediately before bedtime exposure to digital devices and computer screens [LED with low wavelength light in the blue region has been shown to reduce the evening increase of melatonin and tiredness. Additionally, it was discovered that such exposure led to disturbance in nighttime declarative memory and sustained attention performance. (17, 34). The nighttime changes in melatonin concentration and other physiological indications of human biological clocks are suppressed by bright displays. (18).

#### Duration of the computer work

was directly associated to eye symptoms, and prolonged computer use tended to produce complaints that lingered even after the work was finished. (19, 20).

### Symptoms of Computer Vision Syndrome

The most common symptom is eye fatigue: muscle around the eye, like any other, can get tired from continued use. Concentrating on a screen for extended periods can cause concentration difficulties and headaches in the temple and eyes, the Adjectives of symptoms can be distributed into criteria to give the diagnosis of computer vision syndrome (6. 7, 8, 9, 10)

1. **Ophthalmic symptom**:
  A. Visual symptoms - Blurred vision, double vision, presbyopia, slowness of focus change, and Difficulty refocusing the eyes.
  B. Ocular - Internal symptoms- Eye strain, ache in the eye, ache around the eyes, tired eyes, and sore eyes.
  C. Ocular - External symptoms - Burning, dryness, redness, gritty sensation, tearin,g and irritation (13, 14):.
2. **<u>Musculoskeletal symptoms</u>**// Computer workers are at risk of developing Work-related musculoskeletal disorders. These symptoms are well associated with improper placement of computer screens which leads to muscle sprain and include Neck pain, neck stiffness, Shoulder pain, and Backache (6, 7, 8, 9, 10, 11, 16).
3. **<u>Psychosocial symptoms</u>**// psychosocial stress, Fatigue, Dizziness, reduced attention span, poor behavior, irritability, headache, Vertigo // and these symptoms may be related to prolonging of computer use or as a result of exaggeration of Ophthalmic symptoms (1, 14, 15).

**The aim** of the study was to analyze the effects Mobile phones and computers have on eyesight quality in students of the College of Medicine/University of Karbala, who use gadgets for a long time in a day, and analyze the affected of these gadgets in distributed of the computer vision syndrome symptoms

## Subject and Method

This is a cross-sectional study conducted at the University of Kerbala College of Medicine, which was completed between November 2021 and January 2022, to students of the College of Medicine given student questionnaires about the effects of long-time use of computers, laptops, and mobile phone, eyes and different parts of the body, the time spent to collect data was 3 months. The data were collected in the form of online and paper questionnaires. The questionnaire consists of questions about the type of electronic device used for the study, the amount of time spent in front of the device per day, power of eyeglasses if used, symptoms related to electronic devices, and awareness of students about protection techniques

### Subject

The study was conducted on 518 undergraduate students from, the College of Medicine, the University of Karbala with age 19-24 years, consent was taken from them, excluding the samples that did not meet our search criteria and that were about 58 students including the Exclusion criteria which are:

1. The person has an abnormality in vision and used a high degree of eye class more than the power of eyeglasses
2. Disease related to the eye and causes severe headaches as Astigmatism, history of myopia & hyperopia
3. Consumption of drugs that causing affected vision and causes severe headache

### Method

We developed a questionnaire for our subjects, a questionnaire was designed for data collection. The questionnaire has nine subsections dealing with participants’ precise medical history has been taking for each subject, which includes:

1. Identity of the shared subject: name, gender (male or female), date of birth, and grade of the class.
2. What is used in studies and how time use the laptop and mobile
3. Distance more than 50 cm and Using screen filters
4. Medical eyeglasses are used and Degrees of Medical eyeglasses are used
5. Any measures practiced to prevent eye problems
6. Headache
7. Burning sensation
8. Dry tired and sore eyes
9. Double vision and Blurred distance vision of eyes
10. Subject has Neck pain, neck stiffness, shoulder pain and Backache after a long time of use of this laptop and mobile

### Criteria of Computer Vision Syndrome

Three groups of criteria (collection of signs and symptoms in each group), were created in this study required to diagnose the Computer Vision Syndrome, Three signs of these criteria meet and are recurrent in order to diagnose this syndrome, and two of these signs are related to Ophthalmic disturbance and sometimes need treatment, these groups are:

1. Group one// Ophthalmic symptoms
  a. Visual symptoms
  b. Internal Ocular symptoms
  c. External Ocular symptoms
2. Group two// Musculoskeletal symptoms and headache
3. Groups three// Psychosocial symptoms

### Statistical Analysis

was done using Excel and Version 25 of the Statistical Package for the Social Sciences (SPSS). The differences between the groups were evaluated using the correlation and Crosstab to describe the link between the groups, and categorical data were expressed as numbers and percentages Ethical approva

## Result

These cross-section studies do by 33 questions (questionnaires) to 518 students of the College of the Medicine/ University of Karbala, 460 were included in these studies and 58 were excluded according to the criteria recorded in the method. A total of 460 medical students were included in the study with a mean age of (22 ± 1.27). The number of females in this study was 346 students equal to 75.2% and the number of males in this study114 students 24.8%), female with a male ratio of 3:1. Out of 460 students, 164 (35.7%) were using mobile, those were using IPad 158 (34.3%), were using laptops 138 (30%) explained in figure 1.

**Figure1.**
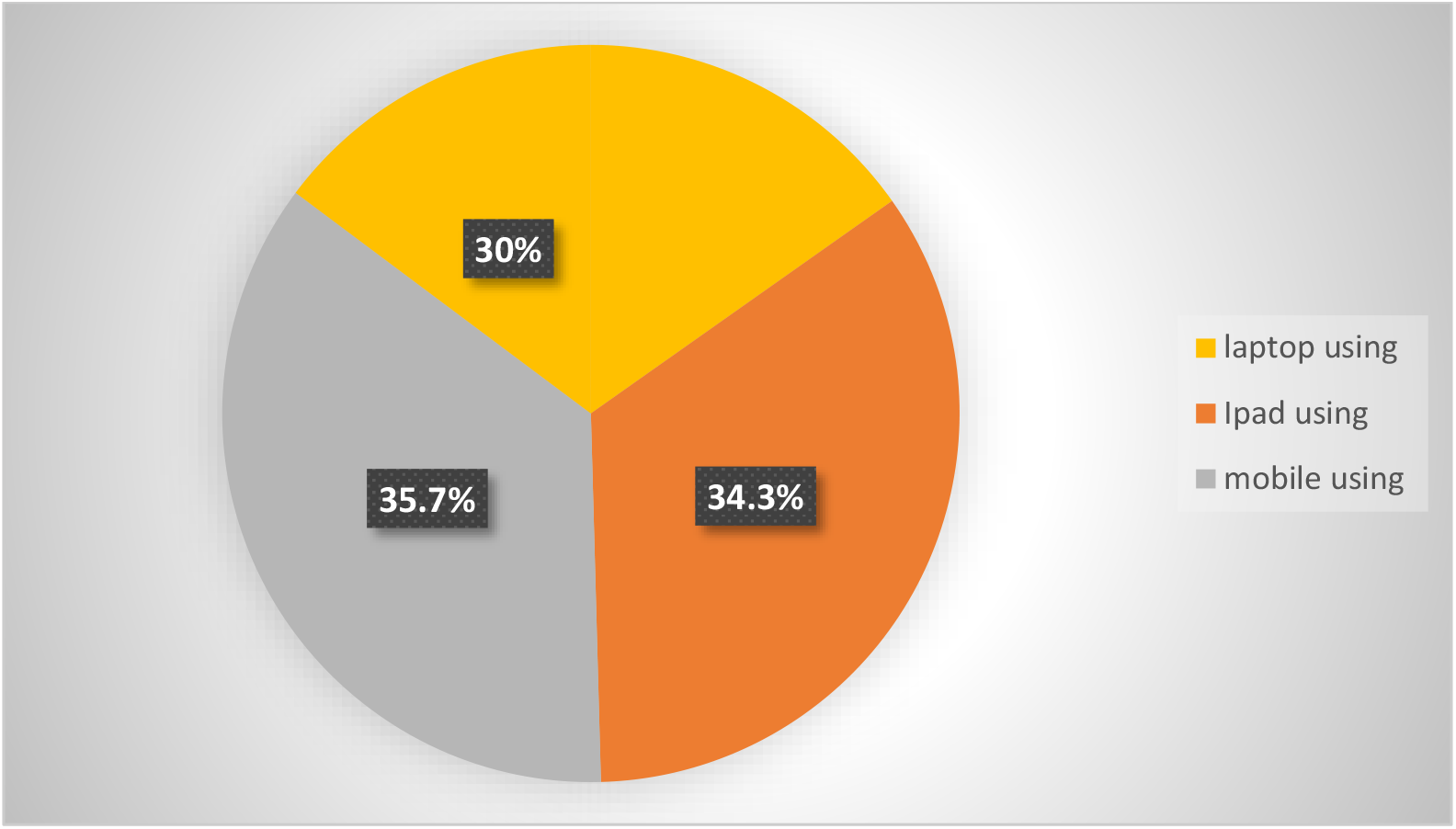
instrument used by the student

The duration of devices use per day for the majority of them was more than 8 hours. Of the students who have used devices from a distance of more than 50cm 228(49.6%) and 232 (50.4%) of them are less than 50cm. 148 (32.2%) of students were using eyeglasses, and the instruments used by students to protect the eyes are explained by the table1.

**Table 1.** the method used for the protective eye from the instrument.

| Preventive measures |  | Frequency | Percent |
| --- | --- | --- | --- |
| Using screen filters | Yes | 156 | 33.9 % |
|  | No | 304 | 66.1 % |
| Taking break | Yes | 278 | 60.4 % |
|  | No | 182 | 39.6 % |
| Illumination of the room | Normal | 344 | 74.8 % |
|  | Abnormal | 116 | 25.2 % |
| Brightness of screen | Normal | 230 | 50.0 % |
|  | Abnormal | 230 | 50.0 % |

The majority of students used the laptop or mobile for more than 8 hours. These cause a problem in vision, Computer vision syndrome prevalence among student participants in this study was 89.6%, of which 70% with the musculoskeletal symptom, the most common vision problem as the ocular symptom reported by the student explained in table 2, and figure 3, and the other problems related to muscular spasms and using devices explained in table 3, and figure 4.

**Table 2.** explains the percentage of symptoms related to vision.

| Type of ocular Symptoms | Total | Number of students with Symptoms related to eye |  |  |  |  |  |  |  |
| --- | --- | --- | --- | --- | --- | --- | --- | --- | --- |
|  |  | Mild |  | Moderate |  | Severe |  | Total |  |
|  |  | Numb<br>er | Percent | Numb<br>er | Percent | Numb<br>er | Percent | Numb<br>er | Percent |
| **Blurred vision** | 460 | 105 | 22.83% | 104 | 22.61% | 29 | 6.3% | 238 | 51.74% |
| **burning sensation** | 460 | 150 | 32.61% | 81 | 17.61% | 39 | 8.48% | 270 | 58.7% |
| **Watery eyes and irritated eyes** | 460 | 178 | 38.7% | 77 | 16.74% | 23 | 5% | 278 | 60.44% |
| **Eye strain** | 460 | 104 | 22.61% | 119 | 25.87% | 47 | 10.22% | 270 | 58.7% |
| **Temporary Double vision** | 460 | 106 | 23.04% | 37 | 8.04% | 9 | 1.96% | 152 | 33.04% |
| **Difficulty in accommodation** | 460 | 93 | 20.22% | 62 | 13.48% | 35 | 7.6% | 190 | 41.3% |

**Table 3.** explains the details of symptoms severity related to musculoskeletal and Extra-Ocular symptom.

| Type of extraocular Symptoms | Total | Number of students with Symptom |  |  |  |  |  |  |  |
| --- | --- | --- | --- | --- | --- | --- | --- | --- | --- |
|  |  | Mild |  | Moderate |  | Severe |  | Total |  |
|  |  | Number | Percent | Number | Percent | Number | Percent | Number | Percent |
| backache | 460 | 132 | 28.69% | 72 | 15.65% | 34 | 7.39% | 238 | 51.74% |
| Shoulder pain | 460 | 101 | 21.96% | 40 | 8.7% | 38 | 8.26% | 179 | 38.9% |
| Headache | 460 | 107 | 23.26% | 134 | 29.13% | 85 | 18.48% | 326 | 70.87% |
| Neck pain | 460 | 62 | 13.48% | 75 | 16.3% | 47 | 10.22% | 184 | 40% |
| Neck stiffness | 460 | 50 | 10.87% | 43 | 9.35% | 11 | 2.391% | 104 | 22.61% |

**Figure2:**
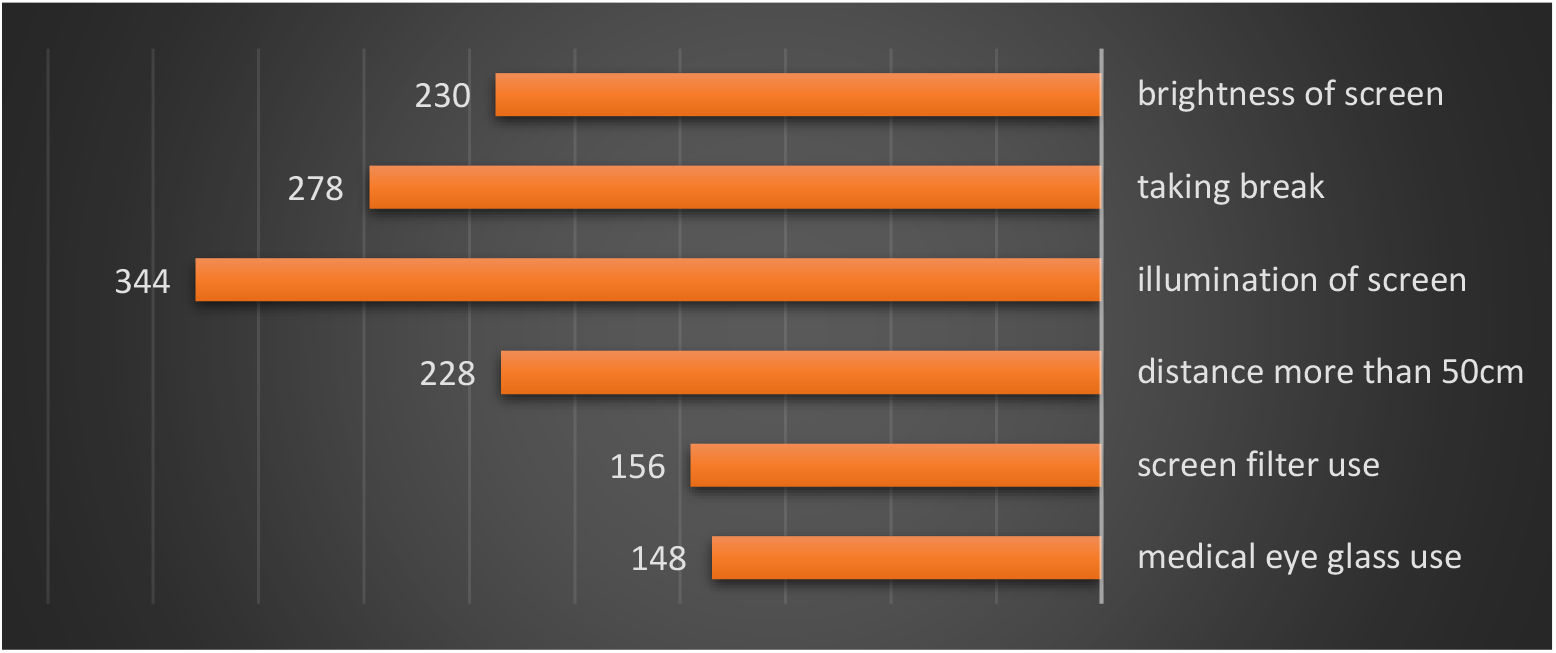
protective eye methods sued by students

**Figure 3:**
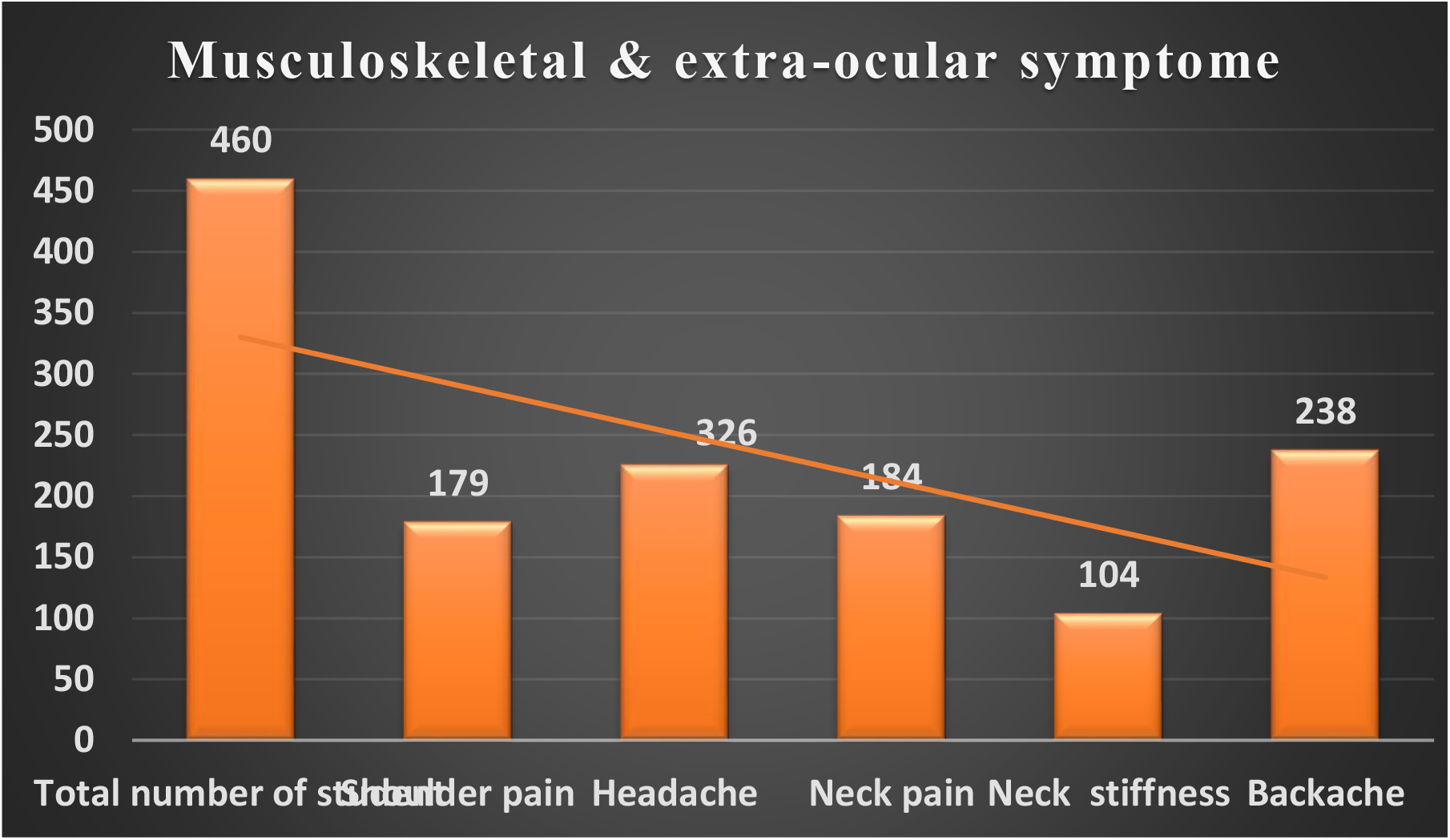
explained the number of students with extraocular symptom

**Figure 4.**
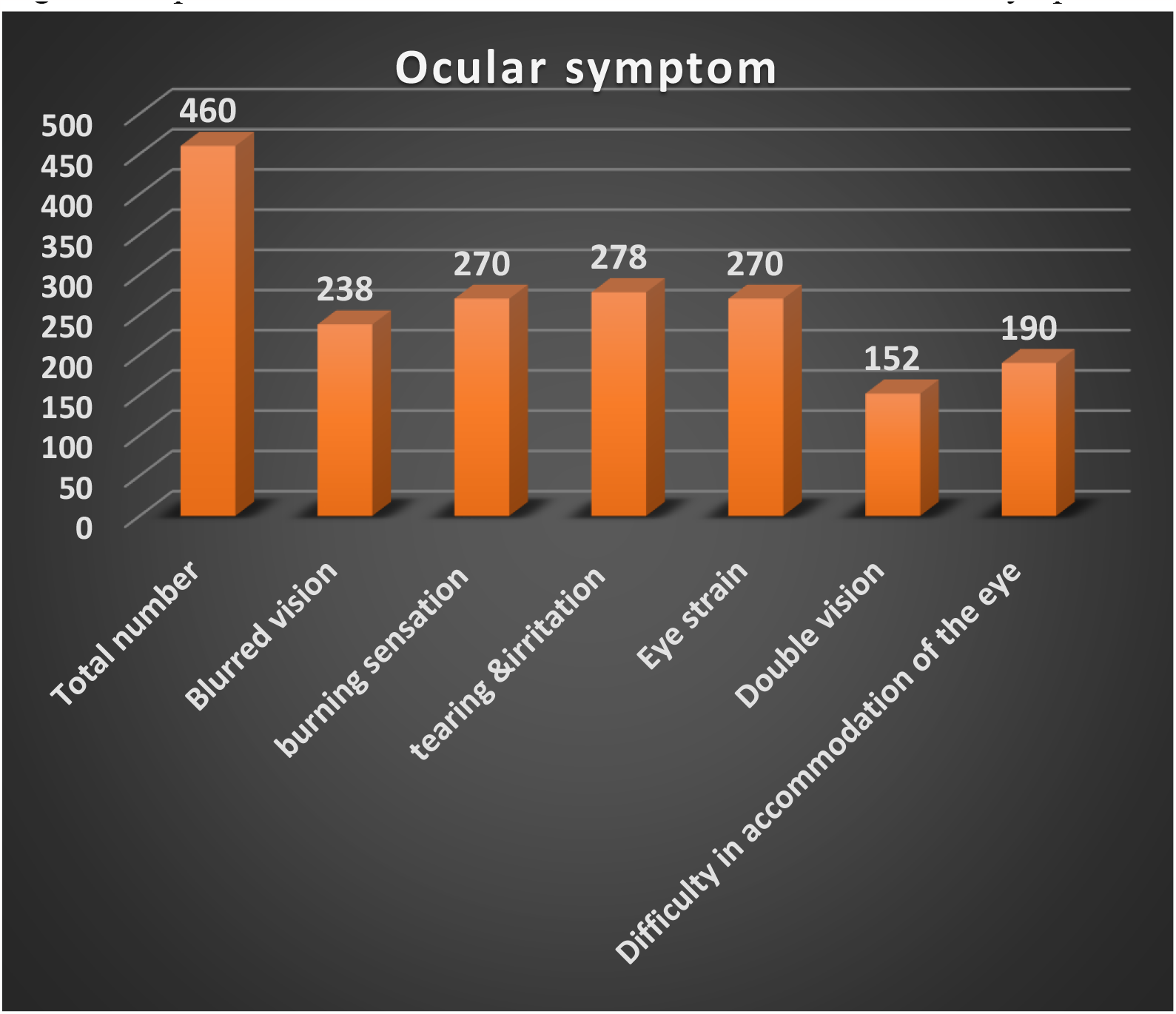
explained the number of students with ocular and visual symptom

To apply the criteria that were created we found 412 of 460 students (89.6% of students) have Computer Vision Syndrome of which 98 students (21.3%) present with the symptoms of all types of criteria recorded with different in severity, well the headache with ocular and visual symptoms in 128 students (27.83%) and backache with ocular and visual symptoms in 140 students (30.43%) and these percentage of the symptoms overlap with each other in some of the cases of students with computer vision syndrome and with other musculoskeletal symptoms.

The Cross tabulation between the tool used by the students’ studies with it and the symptoms of computer vision syndrome in Table 4 and Table 5 explained there are no differences between using the mobile or computer in the number of students with symptoms and the degree of severity of these symptoms

**Table 4.**
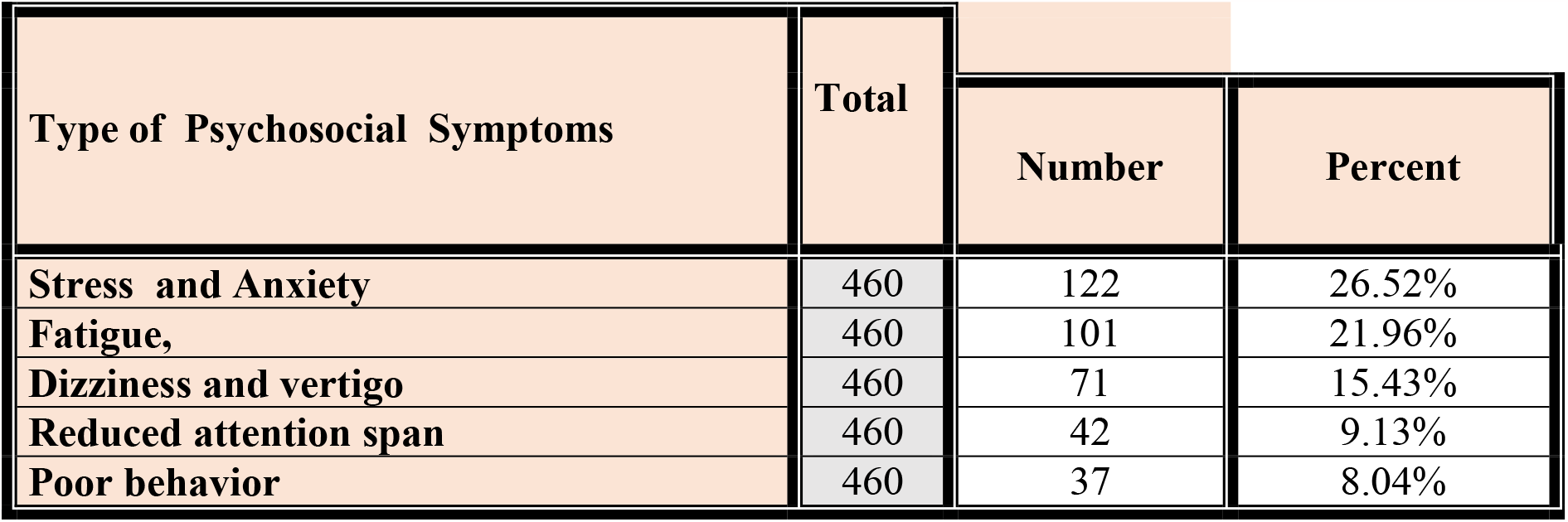
the Psychosocial symptoms distributed in students are explained in Table 4.

**Table 5.** explained the cross-tabulation between the tool used by the students in studies and ocular symptoms.

| symptoms | Degree of severity | The tool used by the students |  |  | Total |
| --- | --- | --- | --- | --- | --- |
|  |  | Computer and laptop | I Pad | Mobile |  |
| **Blurred vision** | Severe | 11 | 8 | 10 | 29 |
|  | Moderate | 30 | 43 | 31 | 104 |
|  | Mild | 32 | 30 | 43 | 105 |
|  | No blurred v | 65 | 77 | 80 | 222 |
|  | Total | 138 | 158 | 164 | 460 |
| **Eye strain** | Severe | 15 | 16 | 16 | 47 |
|  | Moderate | 33 | 49 | 37 | 119 |
|  | Mild | 26 | 28 | 50 | 104 |
|  | No eye strain | 64 | 65 | 61 | 190 |
|  | Total | 138 | 158 | 164 | 460 |
| **burning sensation** | Severe | 9 | 16 | 14 | 39 |
|  | Moderate | 23 | 30 | 28 | 81 |
|  | Mild | 50 | 47 | 53 | 150 |
|  | No burning | 56 | 65 | 69 | 190 |
|  | Total | 138 | 158 | 164 | 460 |

**Table 6.**
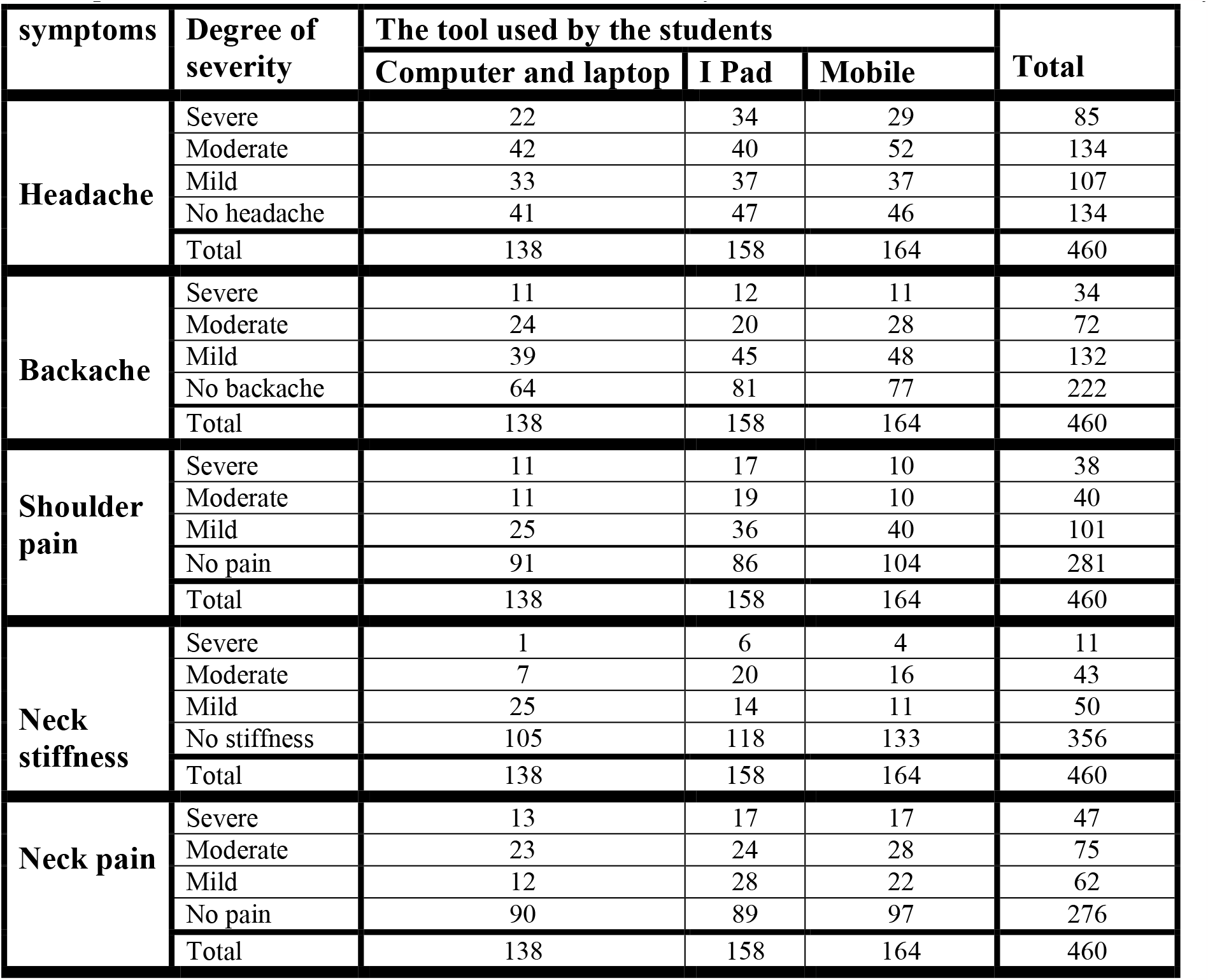
explained the cross-tabulation between the tool used by the students in studies and extra-ocular symptoms.

## Discussion

Midway through October, Iraq experienced a political crisis that resulted in a restriction on college and school enrollment, which was followed by the COVID-19 isolation, which lasted for more than two years and continued teaching, using method of online teaching in schools and universities through online teaching programs and social media has become the result of using mobile phones, iPads, and computers for long hours, and it recorded harm to students in the College of Medicine/University of Kerbala University in addition to covid-19 infection, such as computer vision syndrome. This explains the high prevalence of this syndrome among students, as the combined prevalence rate reached 91.6% compared to other research in other countries did their research before the covid-19 (5, 22, 23, 24, 25, 26) and for certain research during the isolation, the percentage was significantly same percent (27, 28, 29)

Computer vision syndrome results by a person works long hours, uses a computer with an incorrect sitting posture, and does not considerably change the computer’s brightness, Mild symptoms of computer vision syndrome might persist anywhere from a few minutes to several hours while using a computer. When there is a development in cases of severe ocular vision defects, it may be complicated to moderate symptoms that last while using the computer and go away after resting, as well as severe symptoms that require medical attention. Sometimes there is a development in cases of severe ocular vision lesions that leads to the use of lenses and the extraocular vision may be medication.

In this study, we drew lines for the criteria to identify a computer vision syndrome from these associated ocular and extraocular symptoms. in this criteria, the student has ophthalmic symptoms with extraocular symptoms. The ophthalmic symptom common symptoms related to CVS include visual and ocular symptoms, Eye strain, burning, blurred vision, and itchy eyes are the most typical visual issues mentioned in the study. Almost 58.7% of students had eye strain. A lower prevalence in Medical students in Chennai reported a lower frequency of 32.3%, whereas university students in Ajman, United Arab Emirates (UAE), and Jordan reported prevalence rates of 54.8% and 48.3%, respectively (22, 28, 30). Nearly 60.44 percent of our students complained of eye irritation. a lower prevalence recorded among medical students in Chennai, a decreased prevalence of eye irritation (17.4% and 13.9% redness) was noted [22]. While in Ajman, United Arab Emirate (UAE), over 48% of eye irritation cases were reported [30]. In our study, the percentage of Temporary double vision was 33.04%. The lower prevalence was reported in the UAE at 11.5% [30], and in Jordan, it was 18.3% [28]. About 51.71% of our students reported blurred distance vision and this was in accordance with a study done at Al-Aaliyah Amman University in Jordan reported 45.7% [28], in contrast to a study done among medical and engineering students in Chennai was 16.4% and 31.6% respectively [22]. while the prevalence rate of 24.8% was reported in the study that was done in UAE [30], and 48% in the Indian study (32). Difficulty in accommodation percent is 41.3%, This can be explained by the fact that accommodation is a dynamic process and that accommodation fatigue can result from keeping the eyes in a stationary position. By changing the user’s focal point, the user can get relief from constant eye strain and monitor glare (31, 42) Visual fatigue results when the stress caused by these movements on the musculature of the eye exceeds the visual performance ability (10)

Extraocular symptoms may be related to/ or result from musculoskeletal spasms, and accompanied by other ocular symptoms, the most prevalent musculoskeletal symptom experienced by students with computer vision syndrome in this study was backache 51.73%. While reported shoulder pain 38.9% of students, 40% reported neck pain, and 22.61% expressed neck stiffness in contrast to a study done among medical and engineering students in Chennai 60% (22) and in Albania reported a high prevalence of shoulder pain and neck stiffness 81% (33). A study in Jamaica found that 75.1% of undergraduate university students suffered from neck pain and 65.5% from shoulder pain (35). The range is wide because musculoskeletal pain as backache, neck pain and in the shoulder area is influenced by a number of variables, including poor posture and prolonged computer use. But it should be highlighted that this survey shows the highest occurrence, showing the necessity to educate students on proper computer using the right habits.

Headache was the most common symptom reported, 70.87%, In contrast to a study that was done among medical and engineering students in Chennai, about43.3%medical and 45%engineering were reported. Another study was done among university students in Ajman UAE. 53.3% (28), also similar percentage was reported among the university student population in Jordan 53% (30). A similar study in India found 82.1% of the study population reported suffering from headaches (33), Another similar study conducted in Egypt found that 26% of medical students complained of headache (34) common causes of headaches in CVS patients, the additional ocular muscles and ciliary muscles must be constantly contracted to adjust the eyes and keep the lens in the accommodating phase. To see at various distances from the screen to the keyboard and to work on documents, it is necessary to focus and refocus continuously all the time. This causes eye muscle tiredness or eyestrain that causes headaches, in addition to vision-related, there is a cluster headache, and tension headaches—is a common symptom due to frequent computer use.

In our study; Students with Psychosocial symptoms related to computer vision syndrome found high levels of students with Stress and Anxiety at 26.52% and Fatigue at 21.96% compared to those with low levels of Reduced attention span at 9.13% and Poor behavior at 8.04% and these symptoms recorded by other authors in near percent (36, 37, 38, 39, 40, 41, 43). the stress of studies and a political crisis with new online studies Fatigue lead to repetitive feelings of anxiety and stress and continuous feeling of fatigue.

According to our study, students who used iPads and smartphones were more frequently affected by extraocular symptoms of computer vision syndrome than students who used desktops and laptops and had more severity recorded in backache and headache.

## Conclusion

The criteria in this study that can be used to diagnose computer vision syndrome, the severity of the condition, and its relationship to prolonged time of computer or other device use

## Supporting information

Ethical approval

## Data Availability

All data produced in the present work are contained in the manuscript

## ACKNOWLEDGMENT

Many thanks to my students, especially the two fifth-stage medical students (Zahraa Ali and Zainab Ahmed Miri) who assisted me in publishing questionnaires and gathering responses.

## Ethical approval

The study was carried out in accordance with ethical standards established by the Medical Research Bioethical Committee of the University of Kerbala/College of Medicine. Before the start of the questionnaires, informed consent was obtained from the student participants and the study protocol was reviewed and approved by a local ethics committee in number 43 on 24/7/2020. The approval was then renewed annually, and the final approval letter number 83 on 30/7/2023.

