## Supplementary material for "Evaluation of computer vision syndrome in the sample of students from the College of Medicine/University of Kerbala result of online learning with the isolation of COVID-19": Ethical approval

**University of Kerbala**
**College of Medicine**
**Medical Research Bioethical Committee**

### FINAL APPROVAL LETTER

**Mahdy Hameed Abu-Ragheif**
**College of Medicine /University of Kerbala, Dept. of Physiology**
**Date: July, 30, 2023**
**Dear Mahdy**
**Title of Project:**
**Evaluation of computer vision syndrome in the sample of students from the College of Medicine/University of Kerbala result of online learning with the isolation of COVID-19**

**Reference Number:** 83

**Thank you for your recent correspondence. This is to certify that your responses have satisfactorily addressed the research bioethical guidelines. You may now proceed with your research.**
**Please consider the following requirements of approval:**
**1. Approval will be valid for one year. By the end of this period, if the project has been completed, abandoned, discontinued or not commenced for any reason, you are required to announce the Committee.**
**2. However, at the end of the one-year period if the project is still current you should instead submit an application for renewal of the approval. This allows the Committee to fully re-review research in an environment where legislation, guidelines and requirements are continually changing.**
**3. Please remember the Committee must be notified of any alteration to the project.**
**4. You must notify the Committee immediately in the event of any adverse effects on participants or of any unforeseen events that might affect continued ethical acceptability of the project.**
**5. At all times you are responsible for the ethical conduct of your research in accordance with the standard bioethical guidelines.**
**6. The Committee should be notified if you will be applying for or have applied for internal or external funding for the above project.**

**Yours sincerely**

**Assist Prof. Ali Mansoor Al-Ameri**
**Chair, Medical Research Bioethical Committee**
**Date// 30/7/2023**
